# Optimizing delivery strategies for 3HP TB preventive treatment in Tanzania: A qualitative study on acceptability of family approach in HIV care and treatment centers

**DOI:** 10.1101/2024.04.04.24305275

**Authors:** Doreen Pamba, Erica Sanga, Killian Mlalama, Lucas Maganga, Chacha Mangu, Anange Lwilla, Willyhelmina Olomi, Lilian Tina Minja, Issa Sabi, Riziki Kisonga, Emmanuel Matechi, Isaya Jelly, Peter Neema, Anath Rwebembera, Said Aboud, Nyanda Elias Ntinginya

## Abstract

**Introduction:** Tanzania rolled-out a 12-dose, weekly regimen of isoniazid plus rifapentine (3HP) TB preventive treatment in January, 2024. Although 3HP completion rate is generally ≥ 80%, variations exist depending on type of delivery strategy and programmatic setting. Prior to the roll-out, a mixed methods study was conducted to assess whether a family approach involving family member support, SMS reminders and three health education sessions, was acceptable and optimized 3HP uptake and completion. This paper describes acceptability of the family approach among people living with HIV (PLHIV), treatment supporters (TS) and community health workers (CHWs).

**Methods:** This was a qualitative descriptive study in 12 HIV care and treatment centers across six administrative regions. We purposively sampled 20 PLHIV, 12 CHWs for in-depth interviews and 23 TS for three focus group discussions held between September to December, 2023. The theoretical framework of acceptability guided thematic-content analysis using a framework approach.

**Results:** Participants understood that PLHIV have high risk for active TB and that 3HP provides shortened treatment for TB disease prevention. They reported gaining TB and 3HP knowledge from health education sessions. However, participation of TS in health education sessions was low and many reported expensive transportation costs to clinics. Receiving support from someone close and SMS were perceived as good adherence reminders. The majority reported mild self-limiting side effects but expressed positive attitudes because of the shortened treatment, TB counselling, satisfaction from helping others, alignment with lifestyle and work responsibilities and reduced work burden. Some PLHIV reported difficulties in identifying family members for support thus, chose other close friends or CHWs.

**Conclusions:** Delivery of 3HP with support from family members and SMS reminders is widely accepted by CHWs, PLHIVs and TS. Restricting support from only family members was unacceptable and attendance of all three health education sessions by TS may not be feasible.

## Introduction

The co-infection of tuberculosis (TB) and human immunodeficiency virus (HIV) globally remains to be a public health problem and a leading cause of deaths among people living with HIV (PLHIV)(1). Global TB-related deaths among PLHIV were estimated at 190,000 in 2021, a stabilizing trend since 2019 (2). PLHIV are 18 times more likely to become infected with TB and develop active TB than the general population due to the weakened immune system (3). In 2022, PLHIV co-infected with TB contributed to 6.3% of the 10.6 million global TB incident cases (4).

WHO recommends TB preventive treatment (TPT) for prevention of active TB disease among PLHIV, household contacts of bacteriologically confirmed pulmonary TB persons and clinical risk groups such as those receiving dialysis (4,5).The efficacy of available range of TPT regimens is documented to be 60% to 90% (5). The 2018 United Nations High-Level Meeting (UNHLM) on TB committed to expand global TPT coverage to 6 million PLHIV by 2022 (6).Although 11.3 million PLHIV worldwide were reported to have received TPT in 2022 thus exceeding the 2018 UNHLM target of 6 million, a drop of 300,000 PLHIV on TPT was observed between 2021 and 2022 (4).

Tanzania uses isoniazid preventive treatment (IPT) for prevention of active TB disease among PLHIV since 2010 (7). The IPT involves the use of self-administered isoniazid daily for 6 to 9 months (8,9). In 2022, IPT coverage and completion rates were reported to be 84% and 79%, higher than the national coverage and completion targets of 70%and 70%in the same year (10). Countries are recommended by WHO to adopt short course rifamycin-containing TPT regimens (1 to 4 months) that are evidenced to be equally effective, with less hypertoxicity and higher completion rates compared to IPT (5,11,12). The 3HP is among the recommended shorter TPT regimen that involves taking Isoniazid and rifapentine once weekly for 12 weeks. The WHO TPT recommendations of 2020 acknowledge existence of limited evidence on 3HP self-administration (SAT) thus, calling upon additional research including those of digital technologies for adherence support (5). This implies that evidence is needed for novel strategies optimizing delivery of 3HP in routine HIV care settings.

Overall, many studies have documented 3HP completion to be ≥ 80%, higher than SAT IPT in varied TB burden countries under research or routine conditions and have assessed varied 3HP delivery strategies such as; direct observed therapy (DOT), video directly observed therapy (VDOT), SAT alone, SAT with text reminders and facilitated SAT with few conditional clinic visits for DOT (11,13–15). However, 3HP completion rates vary depending on type of delivery strategy and programmatic setting. A study conducted in United States, Spain, Hong Kong and South Africa showed decreasing treatment completion rates with 3HP DOT (87.2%), 3HP SAT with text reminders (76.4%) and 74.0% with 3HP SAT alone (16). Nevertheless, 3HP SAT has better completion rate and is cost-effective compared to IPT (17).

The Tanzanian Ministry of Health had prepared to roll-out 3HP under the guidance of feasibility and acceptability studies (18). Other than DOT, evidence is limited on optimal strategies for delivering 3HP that are feasible and acceptable in Sub-Sahara African programmatic settings. We thus, planned to identify optimized 3HP delivery strategies to support informed decisions on appropriate adherence support strategies during programmatic adoption. We piloted a package of interventions collectively known as a family approach to 3HP to determine acceptability, feasibility and impact on uptake, adherence and completion among PLHIV within the context of a sequential explanatory mixed methods research. As part of the mixed methods research, this paper reports the qualitative component that aimed to assess acceptability of the family approach to 3HP among community health workers (CHWs), PLHIV and their treatment supporters (TS). The full details of the quantitative results will be published separately.

### The family approach to 3HP

This approach involves the use of a package of three 3HP delivery strategies in supporting PLHIV with adherence and completion of 3HP as well as adverse events monitoring. Taken together, the piloted strategies were aimed to improve uptake of 3HP compared to the standard 6 months IPT. They included; the use of a family member identified by PLHIV, automated SMS alerts sent to PLHIV and their treatment supporters and the administration of three TPT health education sessions (enrolment, months 2 and 3). The automated SMS alerts were shared daily for adverse events monitoring, weekly for medication adherence and monthly for clinic visit reminders only to the PLHIV and their treatment supporters who had consented.

The package of interventional strategies was studied in a prospective cohort under implementation research. It was implemented within 12 randomly selected high-volume HIV care and treatment centers (CTCs) of public owned health facilities across 6 purposively selected regions of high HIV prevalence in the country. The cohort formed a quantitative component of a sequential explanatory mixed methods design implemented between May to December 2023. The inclusion criteria for the study participants included; PLHIV enrolled into care within the 12 study CTCs on or after April 2018 to March, 2023 who have identified treatment supporters willing to attend 3 health education sessions as well as those who have completed TB treatment but have not started TPT at the time of study recruitment. PLHIV diagnosed with active TB, with contraindications to rifamycin based drugs or isoniazid, who were already on TPT at the time of study recruitment or were unwilling to use 3HP were excluded from participation. The participants were given 3HP weekly for 3 months, commencing on the 14^th^ day post-HIV diagnosis.

### Theoretical framework

The theoretical framework of acceptability (TFA) was adopted to guide the content of data collection tools, data analysis as well as interpretation of experienced acceptability of the family approach to 3HP. The TFA constitutes seven constructs that inform qualitative assessment of acceptability of health interventions. These include; intervention coherence, self-efficacy, perceived effectiveness, affective attitude, ethicality, burden and opportunity costs(19).The constructs were operationalized to fit the context of this study thus, two of the constructs i.e., burden and opportunity cost were merged and assessed together. We sought to assess the extent to which participants understood TB, TPT, the rationale of the family approach to 3HP/3RH and how it works (intervention coherence), participants’ perceptions of how confident they felt in assuming the roles of treatment supporters/CHWs or in adhering to TPT (self-efficacy), participants’ perceptions of the extent to which the family approach intervention will improve uptake of 3HP (perceived effectiveness), participants’ feelings about being a treatment supporter/ CHW to TPT use or feelings about using TPT (affective attitude), the extent to which the roles of treatment supporters/CHWs or the use of 3HP fits well with lifestyle and work responsibilities (ethicality) as well as efforts and challenges encountered in the implementation of different components of the family approach package (burden/opportunity cost).

## Methods

### Study design

This was a qualitative descriptive study conducted as part of a sequential explanatory mixed methods study. The design enabled us to generate in-depth information regarding perceptions of CHWs, PLHIV and their treatment supporters to understand whether the family approach package to 3HP/3RH was acceptable. Furthermore, it provides additional information that explains results obtained from the quantitative component of the overall mixed methods study that will be published elsewhere.

### Study setting

Qualitative research was conducted in 12 CTCs that were randomly selected to enroll clients into the 3HPTPT across 6 regions in the country namely; Dar es Salaam (Tegeta dispensary and Amana regional referral hospital), Mbeya (Mbalizi designated district hospital and Matundasi dispensary), Iringa (Ngome health center and Mafinga district hospital), Njombe (Njombe town council health center and St. Consolata hospital, Ikonda), Tabora (Tura dispensary and Igunga district hospital) and Mwanza (Misasi health center and Buhongwa dispensary). Permission to conduct the study within the CTC settings was sought from respective health facility authorities.

### Participants and sampling

The study population consisted of three different groups of participants who were purposively sampled namely: PLHIV enrolled into care in selected study CTCs and their treatment supporters as well as supporting CHWs in the respective CTCs. We recruited 20 PLHIV across the 12 study CTCs and who had used 3HP for more than a month. Furthermore, 23 treatment supporters of PLHIV were recruited by HCWs based on their convenience. We maintained a list of CHWs supporting each CTC thus, 12 CHWs from each of the CTCs were recruited.

### Data collection

Data were collected between 14^th^ September 2023 to 9^th^ December 2023 by trained social scientists experienced in qualitative research. Semi-structured in-depth interviews (IDIs) and focus group discussions (FGDs) were adopted as data collection tools. They were consented for audio-recording and were conducted in Swahili for an average of 40 minutes and 90 minutes respectively. A total of 32 IDIs were held whereby; 20 were engaged with PLHIV and 12 with CHWs. Three FGDs of 7 to 8 participants were conducted with 23 treatment supporters of PLHIVs enrolled in 4 CTCs. All IDIs and FGDs were held in the respective CTCs at locations that guaranteed privacy and were convenient for the participants.

The IDI and FGD guides included topics focused around the seven constructs of the TFA in order to inform interpretations on the extent to which the family approach to 3HP/3RH was acceptable to participants. These were composed of questions on knowledge of the family approach package and rationale, confidence in implementing respective roles of the intervention, perceptions regarding the effectiveness of the intervention, attitudes towards the intervention, challenges encountered in use of the intervention as well as perceptions regarding how the intervention aligned with lifestyle and work responsibilities. However, the sample size was determined by the data saturation point during IDI and FGD sessions.

### Data analysis

Three of the six social scientists (DP, ES and KM) engaged in data collection were involved in data analysis. Audio-recordings of IDIs and FGDs were transcribed verbatim and translated into English by respective social scientists who collected the data and who fluent in both English and Kiswahili. We adopted thematic-content analysis using a framework method to analyze the qualitative data. A sample of the translated transcripts from each of the participant groups were exchanged among the social scientists and reviewed to cross-check correctness of verbatim transcription against the audio-recordings by DP & KM. This allowed not only correction of the transcripts but also depth familiarization of the data. Initial versions of codebooks and coding matrices for each participant group were developed by DP based on research questions and the constructs of the TFA. These were later shared to ES and KM whereby revisions were made and agreed as a team. Furthermore, the transcripts were distributed to each of the social scientists for independent coding and charting. Themes were developed based on overarching meanings identified from groups of categories as well as based on grouping TFA constructs interpreted to portray a pattern of similar meaning. The themes were presented in descriptions that included quotations to further illustrate the findings.

### Ethical considerations

Ethical approval was sought from the Mbeya Medical Research and Ethics review Committee (MMREC) in Tanzania (*Ref. No. SZEC-2439/R.A/V.1/168*). Written consent was secured from eligible participants prior to engaging in study discussions. We ensured that the study is conducted in full conformity with the current version of the Declaration of Helsinki and with the Tanzanian health research rules, regulations and guidelines.

## Results

The findings are presented in terms of five themes that relate to the TFA constructs. The these inform our understanding of factors that promoted acceptance of the family approach to 3HP among a majority of participants and those that need to be improved if the approach is to be adopted within programmatic settings.

### Demographic characteristics of study participants

Many of the PLHIV (8/20) were enrolled into care in CTCs from hospitals levels. A great proportion had only primary level education (16/20) and many were earning income from farming (8/20) and small businesses (8/20). They had a median age of 38 years and were living with HIV for median duration of 3.5 months. Majority of the treatment supporters were female (14/23) and many were earning income from small businesses (9/23). Many of them supported PLHIV enrolled into care in CTCs from hospitals (13/23). As for the CHWs, many were volunteering to support CTCs from hospital levels (10/12) and half of them (6/12) had primary level education (See Table 1).

**Table 1:**
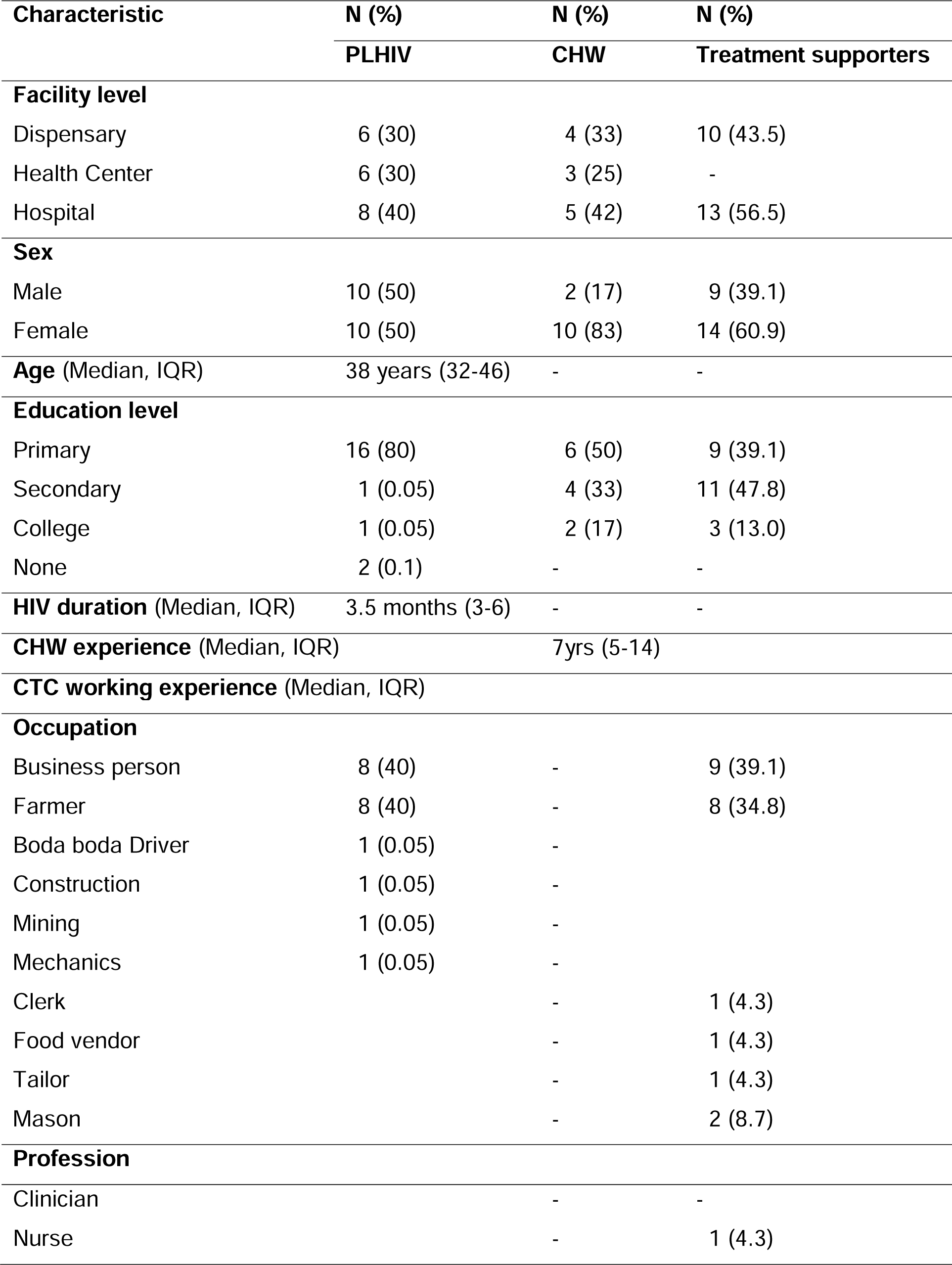
Characteristics of PLHIV, CHWs and treatment supporters.

| Characteristic | N (%) | N (%) | N (%) |
| --- | --- | --- | --- |
|  | PLHIV | CHW | Treatment supporters |
| **Facility level** |  |  |  |
| Dispensary | 6 (30) | 4 (33) | 10 (43.5) |
| Health Center | 6 (30) | 3 (25) | - |
| Hospital | 8 (40) | 5 (42) | 13 (56.5) |
| **Sex** |  |  |  |
| Male | 10 (50) | 2 (17) | 9 (39.1) |
| Female | 10 (50) | 10 (83) | 14 (60.9) |
| **Age** (Median, IQR) | 38 years (32-46) | - | - |
| **Education level** |  |  |  |
| Primary | 16 (80) | 6 (50) | 9 (39.1) |
| Secondary | 1 (0.05) | 4 (33) | 11 (47.8) |
| College | 1 (0.05) | 2 (17) | 3 (13.0) |
| None | 2 (0.1) | - | - |
| **HIV duration** (Median, IQR) | 3.5 months (3-6) | - | - |
| **CHW experience** (Median, IQR) |  | 7yrs (5-14) |  |
| **CTC working experience** (Median, IQR) |  |  |  |
| **Occupation** |  |  |  |
| Business person | 8 (40) | - | 9 (39.1) |
| Farmer | 8 (40) | - | 8 (34.8) |
| Boda boda Driver | 1 (0.05) | - |  |
| Construction | 1 (0.05) | - |  |
| Mining | 1 (0.05) | - |  |
| Mechanics | 1 (0.05) | - |  |
| Clerk |  | - | 1 (4.3) |
| Food vendor |  | - | 1 (4.3) |
| Tailor |  | - | 1 (4.3) |
| Mason |  | - | 2 (8.7) |
| **Profession** |  |  |  |
| Clinician |  | - | - |
| Nurse |  | - | 1 (4.3) |

### Theme 1: Knowledge about family approach to 3HP (intervention coherence)

This theme describes the extent to which CHWs, PLHIV and their treatment supporters understood the rationale of the family approach to TPT and their general understanding of TB and TPT. It relates to the TFA construct of intervention coherence.

Many of the PLHIV understood that TB is transmitted through the air and in close proximity to someone who is sick from TB. They reported being educated by HCWs at the time they were diagnosed with HIV. They all knew that they are at risk for TB because of low immunity caused by HIV.

> “*According to what I was taught here at the clinic, PLHIV are at high risk of acquiring TB because our immunity is already low*…” (Female PLHIV, IDI, dispensary).

They reported taking 3HP once weekly for three months and understood that TPT is to help them from acquiring TB disease but were unable to narrate details about TB treatment. Few reported being informed of long and short TPT regimens. The quote below narrates a client’s understanding of why she was given TPT:

> “*I was told by the nurses who gave me those medication, they said that since I am HIV positive then my immunity is low so I can be attacked by any disease any time, that’s why they gave me TB preventive treatment*.” (Female PLHIV, IDI, dispensary)

Furthermore, the PLHIV understood that the family member is someone to support them in medication adherence. They understood that a treatment supporter should be someone close thus, many reported selecting their spouses. These conceptions are illustrated in the quotes below:

> “*[treatment supporter should] tell him or her that when it reaches certain hours, have you taken the medicine or not?*” (Male PLHIV, IDI, dispensary)

> “*They told me to choose my closest person…the close person I have here is my husband, I can’t go to another house to find the treatment supporter, that’s not possible and that’s why I preferred my husband*” (Female PLHIV, IDI, health center)

However, one of the PLHIV did not see the need or importance of having a treatment supporter as he perceived himself as able to take care of his own health and not needing supervision from someone else

> “*Personally, I just can’t say that you need someone to supervise you so that you can take the medicine because this is your own health and when you will take the medicine that means you are protecting yourself, you just can’t say that until someone supervises you that’s when you will take the medication, I am taking control of my own health so that I can be okay, I don’t need anyone to observe me*” (Female PLHIV, IDI, health center)

Similarly, many of the treatment supporters understood that TB can be transmitted through the air and from someone who hasn’t started TB treatment. All of them said that TB is treatable and cured within 6 months of treatment. They mentioned their clients to be at risk for TB disease because of low immunity from HIV and having being educated by HCWs that TPT prevents TB among PLHIV. This is illustrated in the quote below:

> “…*the preventive treatment is for the person who is not infected with TB. Therefore, before he was given those drugs, he was screened for TB by being asked for TB signs and symptoms where among the questions I remember one was about coughing. After that the doctor said if a person is found with TB cannot use preventive therapy and instead can use TB treatment drugs for six months*” (Male Treatment supporter, FGD, dispensary)

The treatment supporters understood that the family approach intends to help the client at a family level. They knew that as a treatment supporter, they are supposed to remind their clients about taking medications and also identification and reporting of TPT side effects.

> “*Sometimes, the patient may not adhere to the treatment. By reminding them, they can take their medication without hesitation or quitting. If they encounter any challenges, they can seek advice from the treatment supporter before going to the healthcare provider*” (Female Treatment supporter, FGD, hospital)

> “…*as per doctor’s instructions it seems my presence to this patient is needed because he might get those effects to the extent that he cannot be able to report, that makes me as part of the family to report the incidence to the health facility that my relative has got such effects for them to assist the patient*” (Male Treatment supporter, FGD, dispensary)

All of the CHWs understood that 3HP is a shortened TPT compared to INH and is given to someone confirmed of not having TB/TB symptoms but at risk for TB. They stated that PLHIV have freedom to choose the day and time to take 3HP.

> “*It is taken for three months, once in a week and the client will choose the specific day to take them for example he can choose Wednesday, so every Wednesday he will be taking them till three months*” (Female CHW, IDI, health center)

The CHWs understood that the family approach intends to use a family member to support TPT adherence which will in turn also protect the family from contracting TB.

> “…*as you are teaching them sometimes the client might not concentrate due to his condition so the treatment supporter will help to remind him that at the hospital, we were told about this and that. For example, the client might decide to stop taking the medication saying that he is tired but the treatment supporter will intervene to tell him that we were insisted that you can’t skip any day, so just take them, also he will encourage him*” (Female CHW, IDI, health center)

However, they reported that a closed one can assume the role of a treatment supporter not necessarily a family member.

> “*We ask the client to suggest the person he thinks he will be comfortable with like who do you think you can disclose to? is it your father, mother, your child, a friend or just any one?*” (Male CHW, IDI, hospital)

### Theme 2: Confidence in 3HP adherence (self-efficacy)

This theme describes the treatment supporter’s confidence that he or she can support his or her client to adhere to TPT. It also describes the confidence of PLHIV in adhering to TPT. As for CHWs and in the context of family approach, it refers to the CHW’s confidence that he or she can support his or her client to adhere to TPT and conduct household HIV testing. The theme relates to the TFA construct on self-efficacy.

The PLHIV were confident in their ability to adhere to 3HP and clinic visits because they understood that they were at risk for TB.

> “*I knew that I have already got this [HIV] disease therefore I have low immunity, so, I better take the [TB] preventive medicines so as when the [TB] disease enters at least find me prevented already*” (Male PLHIV, IDI, dispensary)

All treatment supporters reported compelling reasons that influenced their acceptance of offering support. Many of those who were first degree relatives reported an obligation to provide support to the clients based on blood relations and dependability. This was also the case for those who were close friends.

> “*It was my responsibility to bring my mother to the hospital for treatment when it was discovered that she had HIV. So, she needed immune protection to stay healthy. I had no fear because I knew it’s all part of life’s challenges, and she needed my support.*” (Female Treatment supporter, FGD, hospital)

> “*In my case, it’s my sister who went through this ordeal, and I wasn’t advised by anyone. But I thought it was better to be together and provide encouragement because life unfolds this way. If I had left her alone, she might have suffered psychologically.*” (Female Treatment supporter, FGD, hospital)

One of the treatment supporters was a CHW who stated having empathy for PLHIV who did not have someone close to support him or her thus, being willing to assume the role of a treatment supporter.

> “*It is when you see people losing their lives, they are suffering because of not having a helper or an adviser; that is when I was motivated that whenever it such kind of a person happens, I must support him/her…I must encourage him so that he uses the medicine to save his life, so that other things in life can proceed*” (Female Treatment supporter, FGD, hospital)

Few of the treatment supporters reported feeling motivated to continue supporting treatment because of having clients who remembered taking the drugs on their own prior to being followed up.

> “ *The patient himself has made me confident to continue supporting him well he is the one who showed me cooperation by taking medications, he does not afraid of taking pills only that my responsibility is to remind him on the day and time to take medicines*” (Male Treatment supporter, FGD, dispensary)

All of the CHWs felt confident of being able to conduct household HIV counseling and testing because of numerous trainings that they received as well as being known in their communities.

> “*I can say that I have confidence in myself since I have been doing this counseling for a very long time, the more I continue talking to people and various facilities the more I gain confidence in counseling, some are even coming to my house so that I can offer to them counseling.*” (Female CHW, IDI, hospital)

> “…*I trust my ability because when you want to approach the household, you just can’t get there and say I have to test on you, no. When you get there, you have to explain why you went there, the advantages and disadvantages, after narrowing those down then he will understand*” (Female CHW, IDI, dispensary).

### Theme 3: Perceived effectiveness of family approach (perceived effectiveness)

The TFA construct on perceived effectiveness was linked to this theme that describes participant perceptions of the extent to which the family approach will increase uptake of TPT.

The PLHIV viewed using a closed one as a treatment supporter is very helpful to support their 3HP adherence. They narrated receiving varied advice, medication reminders and support for food from those whom they selected as their treatment supporters. Many of the PLHIV also mentioned having being given the freedom to choose anyone to support them based on closed relations.

> “*We normally have discussion with him, he even advises me on what I should eat, type of work I should do, sometimes he even sends the texts to remind me on different issues related with TPT*” (Male PLHIV, IDI, dispensary)

> “…*You can choose your relative or your close friend who can keep your secret*” (Male PLHIV, IDI, hospital)

The PLHIV viewed using a closed one as a treatment supporter is very helpful to support their 3HP adherence. They narrated receiving varied advice, medication reminders and support for food from those whom they selected as their treatment supporters. Many of the PLHIV also mentioned having being given the freedom to choose anyone to support them based on closed relations.

> “*We normally have discussion with him [treatment supporter], he even advises me on what I should eat, type of work I should do, sometimes he even sends the texts to remind me on different issues related with TPT*”(Male cPLHIV, IDI, dispensary)

> “…*You can choose your relative or your close friend who can keep your secret*” (Male PLHIV, IDI, hospital)

They commended on how the SMS texts reminded them to take their pills. Few stated how they felt cared for by the HCWs upon receiving the texts.

> “*The texts are really helping me…Sometimes I get busy and I might forget to take them, but just a short time before my time I would receive the text, So I know it’s time*” (Female PLHIV, IDI, dispensary)

> “*I enjoy receiving those texts because I feel that you are caring about me, and when I receive a text, I become cautious that I am supposed to do something.*”(Male PLHIV, IDI, dispensary)

All of the PLHIV perceived the health education sessions to have increased their knowledge of TB and TPT. One of the PLHIV stated being influenced to be very careful in preventing oneself from surroundings that would increase his risk for TB transmission.

> “*The first advantage is that they are giving you the education that you didn’t know before, and if you really pay attention to them, it helps you…For example, I have learned how to protect myself like taking precautions when one is coughing, maintaining some distance or when you are in a group of people how should you sneeze and other things, so I am paying attention to those details*” (Male PLHIV, IDI, hospital)

The treatment supporters perceived the use of automated SMS as a good reminder system for medication intake as the drugs are taken weekly thus, easy to forget.

> “*It is not always for the human mind you will be thinking about when the patient will take medicine, but when I see that message on that day, I do not forget I tell her*” (Female Treatment supporter, FGD, health center)

They stated that health education sessions did not only improve their client’s health but also the community as a whole.

> “*To me, health education helps me in this time and later, because if I will be able to let this one not get TB, that means I will be able to support another person, be in the family or outside the family which is the general community*” (Female Treatment supporter, FGD, health center)

One of the treatment supporters perceived the one-to-one health education sessions as challenging and recommended that they are conducted among groups of clients and their treatment supporters.

> “…*I don’t know if they have a system to gather patients together. That’s what I consider a hindrance because each patient arrives at different times due to their different appointments.*”(Female Treatment supporter, FGD, hospital)

All of the CHWs viewed the automated SMS as good reminders for timely intake of drugs. However, one of them was concerned of clients who were unwilling to receive the texts.

> “*About the SMS, I think it is a good thing because if the client is taking the medication once in a week, then it is easier to forget but once he receives the TPT text then it is easier to remember or even if the Treatment supporter receives the text, then it is easier for him to remind him*…” (Male CHW, IDI, dispensary)

> “*There are challenges because some of clients at the beginning refused to be sending them these texts…when you ask him, he would say don’t send them I will remember on my own…He is worried that someone else might see that text*” (Female CHW, IDI, dispensary)

### Theme 4: Attitudes towards family approach (affective attitude and ethicality)

This theme describes feelings about using TPT or feelings about being a supporter for TPT use in the context of the family approach. It relates to the TFA constructs on affective attitude and ethicality.

The PLHIV expressed positive attitudes about the different components of the family approach to 3HP. These were attributed to the fact that 3HP is shorter than INH and it prevents them from TB.

> “*When I heard that this medicine will protect you against acquiring TB and I have already witnessed how TB patient is suffering, so I got motivated and I was very happy*…” (Female PLHIV, IDI, dispensary).

Others felt happy about having a treatment supporter and the counseling they received about TB and TPT at the clinics.

> “*I thought one day I might get sick to the point that I can’t leave my house or even take a glass of water, then by having a treatment supporter he can help me to come to collect my medication and support me with other issues*”(Male PLHIV, IDI, dispensary)

Treatment supporters perceived their role as a blessing to serve people’s lives. One of them stated that even the clients consider them as their confidants and supporters to improve not only physical but also psycho-social well-being intended for the patient’s health and welfare.

On the other hand, few treatment supporters mentioned the issue of self-patient stigma against their supporters in the sense that some patients did not accept the supporter to accompany them to the health care facility though they are positive with phone follow-up.

> “*My client, let us say it is about stigma…he said I cannot go with a person to the hospital because I will be seen that I am taken to the hospital. Just take my contacts…But since he is already into dose, I did that and he was faithful he went for the service and he is continuing with the services well…I have never attended with him and he does not want* “(Female Treatment supporter, FGD, dispensary)

CHWs also expressed positive attitudes towards the family approach to TPT. They reported liking 3HP as it is a shorter TPT and also liked the use of a treatment supporter who in turn simplifies their work.

> “*I would like to thank you about TPT and most importantly our clients were taking a six months dose but now within just three months they have already completed their dose, and before they used to take them on daily basis but now it is just once in a week, so I am very grateful that TPT is just within a short period of time, the client isn’t getting tired*” (Female CHW, IDI, dispensary)

> “*I really like this aspect because it is reducing some of my burdens and it makes my work to be effective because if I miss my client and he has no treatment supporter then not becomes very difficult for me to locate him, but if he has a treatment supporter…it is easier to reach the client through [him]*…” (Male CHW, IDI, hospital)

We sought to explore how the role of treatment supporter in the TPT study or the role of CHW within family approach to TPT fits with their daily lifestyle and work responsibilities. This also applied to how the use of TPT fits well with PLHIV lifestyle and work responsibilities.

There were no changes in lifestyle or work responsibilities that were reported as a result of taking 3HP. One of the PLHIV commented on how free he felt to take the drugs in front of other people.

> “*No, it hasn’t changed anything because first I can take my medication even in front of other people since not so many people knows about those medication, when it is time even if there are guests at my house, I will just take my medication, if they ask I can it is just the ulcers. I can say many people knows the ARV but not TPT medication*.” (Female PLHIV, IDI, dispensary)

Majority reported not having experienced lifestyle changes as a result of being a treatment supporter. However, one of the treatment supporters stated not having the time to converse with his patient about his treatment progress due to being busy with work.

> “…*you wish you could spend more time with the patient to understand their challenges better. However, I am an entrepreneur, and in order to have a conversation with the patient and get to know their challenges, I have to leave for my business. You may plan to visit your patient today, but you find that you are running out of time to be at home with your patient and understand their challenges because you are late returning from your business*.” (Male Treatment supporter, FGD, hospital).

Few others complained how they had to leave work or lost their job because their client was sick and had to be taken cared for.

> “*The challenge I face is from the moment my patient started getting sick until now. It requires me to be available all the time due to our temporary jobs. We can’t rest even during the weekend, so I ended up losing my job*.” (Female Treatment supporter, FGD, hospital)

Similar to PLHIV, no changes in work responsibilities were reported by CHWs because they viewed that the changes did not relate to an additional work but rather change in type of TPT education they had to provide.

> “*Nothing has changed since this is the same thing we have been doing, the only thing that has increased is providing education about TPT, they are the same responsibilities we have been fulfilling because when we visit a PLHIV we must perform TB screening, and these two things are going together. When I visit him, I will speak about HIV, about TB screening as well as TPT*” (Female CHW, IDI, hospital)

### Theme 5: Difficulties in implementing family approach to 3HP (burden and opportunity cost)

The two TFA constructs on burden and opportunity costs informed the development of this theme that describes efforts and challenges encountered during the implementation of different aspects of the family approach to 3HP/3RH provision.

Few of the PLHIV reported difficulties in selecting someone close to support them in TPT because they feared disclosing their status to their relatives and perceived not to have close friends who could keep a secret.

> “*Till now still I haven’t selected any [treatment supporter] because I am living on my own, my relatives are very far…first since I was diagnosed positive, I decided to stay away from my friends…I do have friends but a close one whom I can tell my secret*…” (Male PLHIV, IDI, hospital)

Many reported experiencing drug side effects especially during the first month but were tolerable. These include; reddish urine, nausea, fatigue and headache. One of the PLHIV stated reaching out to his treatment supporter for advice as narrated in the quote below:

> “…*I was discharging a reddish urine, I even called my sister to ask her about it, she told me that “remember now you are taking several medications, so you should be ready to accept any changes*” (Female PLHIV, IDI, dispensary)

When probed about 3HP adherence challenges, treatment supporters reported patient complaints about drug side effects such as nausea, fatigue and reddish urine. Additional challenges included; not having enough money to support themselves or their clients with transport fare to the clinic and for food costs, not being able to spend quality time with their clients as well as not adhering to the exact time of taking their drugs. These are illustrated in the quotes below;

> “…*when my client started using them, he started having colored urine…I also started being afraid because I thought it was blood, but later on I asked the providers and they said no…he is progressing well*” (Male Treatment supporter, FGD hospital)

> “…*because we are living a bit far from here…therefore, from there up to here and back sometimes is difficult because I lack money for transport*” (Treatment supporter, dispensary)

In regards to participation in health education sessions, treatment supporters commented on their ability to support their clients during the first sessions but one of them felt that they were rushed so that HCWs could accommodate long queues at the clinic.

> “…*when I went there, the health education sessions, we felt like they were rushed. They had many customers to attend to and didn’t have time to explain properly*” (Female Treatment supporter, FGD dispensary)

They stated a tendency of their clients to arrive at the clinics late contrary to the agreed meeting time thus, making it difficult for them to manage the clinic visit delays. Moreover, they perceived their attendance to second and third sessions as unnecessary as the information could be relayed to them by their clients. This is narrated in the below quote;

> “*Regarding health education…we were together me and my client on the first day, and that situation depends on relationship that you have built with your client…your client will just go and bring explanations on what was said…you are called to come and there is long queue it becomes difficult*” (Male Treatment supporter, IDI, hospital)

CHWs stated that some of the PLHIV were unwilling to receive SMS alerts and some who lived in remote areas were unable to receive the SMS texts timely as narrated in the below quote:

> “*When they are in remote areas that means they have poor network and they won’t receive the texts on time…when he returns to refill and you ask him if he has been receiving the texts, he would say no while others who live around town, they get them*” (Male CHW, IDI, hospital)

They reported challenges that they anticipate to occur when 3HP is scaled up for programmatic use. These included difficulties in receipt of automated SMS reminders for clients who; do not own mobile phones, reside in rural areas with network connection problems, have a tendency of not reading text messages or are unwilling to accept receipt of SMS alerts.

> “*The challenge that I see maybe the client who is using a relative’s phone, you know the relative isn’t always around, sometimes he is away on his busy schedule, he might receive it today but he will call tomorrow to inform him or he might neglect it, so the client won’t get it as a reminder*.” (Female CHW, IDI, health center)

## Discussion

This study sought to assess acceptability of a family approach to delivering 3HP among PLHIV, TS and CHWs. According to TFA, an acceptable intervention is that which beneficiaries consider it as “appropriate and satisfactory, based on anticipated or experienced cognitive and emotional responses to the intervention” (19).

The findings indicate that participants were knowledgeable about vulnerability to TB, TPT and family approach. They perceived the approach as important for supporting 3HP adherence and completion and believed that they could adhere and complete treatment given their risk for developing TB. Although a majority of PLHIV experienced mild self-limiting side effects, they had positive attitudes towards two components of the family approach (i.e., having a family member and SMS reminders). This shows that receiving support from a family member and SMS reminders was acceptable to the majority. Perceived TB risk is also reported as a facilitator for 3HP acceptance in other studies in Africa(20,21).

WHO advocates use of digital technologies for attaining the End TB strategy 2035 targets(22) and studies have shown that digital technologies enhance latent TB treatment adherence and is acceptable to PLHIV(13,21,23). Reminder SMS could thus, be a good option to improve 3HP adherence and attain desired completion rate among PLHIV. However, caution is to be taken to ensure that confidentiality is preserved to avoid unintentional HIV status disclosure.

We observed challenges among a minority of PLHIV who due to varied reasons were unwilling or unable to identify a family member for treatment support, were unwilling to receive SMS and a majority of treatment supporters who were unable to attend health education together with their clients. These imply that the family approach is not a one size fits all strategy and that diverse needs of PLHIV have to be considered. The family approach may be adopted as a patient-centered approach to 3HP provision where PLHIV who are unable to receive treatment support from family members can opt for other significant others or receive SMS reminders. Provision of patient-centered TPT services within differentiated service delivery models is called upon for improved TPT coverage and increased completion rates(24–27)and is emphasized as the first pillar by the WHO End TB strategy for achieving End TB targets (28) thus, customizing each of the family approach delivery components to PLHIV preferences is expected to enhance 3HP adherence and completion.

Low participation of treatment supporters in second and third heath education sessions indicate that this component of the family approach does not align with their values and lifestyles thus, is unacceptable. Furthermore, treatment supporters’ perception that participation was expensive and feelings that the sessions were rushed with HCWs due to work burden might have influenced negative attitudes thus, discouraging participation. This may influence the family approach to be generally ineffective given receipt of limited training and counseling. Studies have indicated the effectiveness of family involvement in supporting compliance to TB prevention and treatment when they have positive attitudes and sufficient training (29–31).

Although family approach is shown to be acceptable to participants, the qualitative design cannot provide conclusive arguments that it is effective in increasing 3HP adherence and completion. Quantitative findings of the impact of family on 3HP uptake and completion will be published separately.

## Conclusions

Delivery of 3HP with support from family members and SMS reminders is widely accepted by CHWs, PLHIVs and TS. However, the family approach is not a one size fits all strategy thus, is recommended as a person-centered approach for 3HP delivery among PLHIV. Restricting support from only family members was unacceptable and attendance of all three health education sessions by treatment supporters may not be feasible. Other relatively cheap platforms to engage and educate family members are needed if the component on family involvement is to be adopted in programmatic settings.

## Supporting information

In-depth interview guides

## Data Availability

Study participants were assured of data confidentiality when securing informed consent. Some of the interview transcripts contain identifiable information thus, making raw data available will be a breach to confidentiality. The data that support the study findings are available from the corresponding author upon reasonable request. Please contact for data availability requests.

## Acknowledgements

The authors are thankful for all participants who consented for discussions in the study. We extend our gratitude to Nasra Abdul, Simeon Mwanyonga and Lonze Ndelwa for supporting data collection and transcriptions. We commend the support received from Dr Catherine Joachim on behalf of the Tanzania Ministry of Health for ensuring a supportive environment for implementation of the study.

## Author contributions

Study conceptualization: D.P., E.S., Data collection and analysis: D.P., E.S., K.M. Wrote the first draft of the manuscript: D.P. Writing (critical review and editing): D.P., E.S, K.M., L.M., C.M., A.N., W.O., R.K., I.S., A.R., S.A., N.E.N., All authors have read and approved the final manuscript.

## Supporting Information

S1: In-depth interview guide_people living with HIV

S2: In-depth interview guide_community health workers S3: Topic guide_treatment supporters

