## Supplementary material for "Optimizing delivery strategies for 3HP TB preventive treatment in Tanzania: A qualitative study on acceptability of family approach in HIV care and treatment centers": In-depth interview guides: S1_In-depth interview guide_People living with HIV.docx

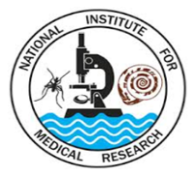
 **IN-DEPTH INTERVIEW GUIDE (PLHIV): English**

**Interviewer Information**

Name of Interviewer:

Location:

Interviewee code:

Date:

**Introductory Remarks:**

Thank you for consenting to participate in this in-depth interview. I am.......................... and I would like to discuss with you about your views on the use of Tuberculosis preventive treatment (TPT) as prophylaxis against TB disease. In particular, I would like to understand the challenges you have experienced and ways you have been able to cope with them.

**Discussion Topics**

1. I would like to start by collecting some demographic information: sex, age, education level, occupation and duration of HIV

**Knowledge about family approach to TPT provision (Intervention coherence)**

2. Please tell me what you understand about HIV
   - *transmission*
   - *high risk groups*
   - *treatment*
3. Please tell me what you understand about Tuberculosis

- *transmission*
- *high risk groups*
- *treatment*
- *prevention (prophylaxis)*

4. What do you understand about Tuberculosis preventive treatment?

- *uses of TPT*
- *duration*
- *who should take it?*
- *dosing, accessibility*
- *what do others say about it?*

5. Please tell me what you know about using a family approach to TPT provision

- *rationale*
- *procedures involved*
- *how was information obtained?*
- *stakeholders involved*

**Confidence in performing the behavior required for family approach to TPT**

**(Self-efficacy)**

6. What motivated you to accept taking daily TPT via the family approach?

- *TB risk perception, influence from others*

**Capacity of family approach to increase uptake of TPT (Perceived effectiveness)**

7. In your opinion, what can hinder the ability of using treatment supporters to support the use of TPT among PLHIV?

- *busy lifestyle*
- *impatience*
- *uncommitted*
- *lack of support skills*

8. How has daily SMS reminders supported you in taking TPT and in identifying TB symptoms?

- *adherence, recognition of TB symptoms*

9. What barriers did the community health worker experience when reaching out for your household members for HIV testing and TB screening?

- *unwillingness of household members*
- *privacy issues*

10. What could hinder health providers to provide better services for TPT during clinic consultations?

- *patient workload*
- *unfriendly language*
- *inadequate counselling (privacy)*

**Feelings about receipt of TPT via family approach (Affective attitude)**

11. Please tell me about your treatment supporter

- *who and why him/her?*
- *how is support given?*
- *challenges (coping strategies)*
- *level of commitment for support*

12. Please tell me about your community health worker

- *what aspects do you like about his work?*
- *what aspects of his work need improvement?*

13. When you think about the way in which TPT has been offered to you, from how you were approached to the services you have been receiving so far, what aspects do you like?

- *counselling*
- *use of treatment supporters*
- *support from CHW*

14. When you think about the way in which TPT has been offered to you, from how you were approached to the services you have been receiving so far, what are the aspects that you do not like?

- *counselling*
- *side effects*
- *clinic visits*
- *use of treatment supporters*
- *support from CHW*

**Intervention fitness with lifestyle and work responsibilities (Ethicality)**

15. Please tell me how you take your TPT?

- *timing*
- *storage*
- *disclosure of use*
- *challenges and coping strategies*

16. In your opinion, has daily TPT influenced changes in the way you live?

- *changes in work schedule, anxiety*

**Efforts/challenges experienced in using TPT via the family approach (Burden/opportunity costs)**

17. How has your treatment supporter supported you in taking TPT?

- *phone calls and text messages*
- *direct observation*
- *counselling*
- *challenges the two of you have experienced, coping strategies*

18. What opinions do you have about participating in health education sessions at the clinic with your treatment supporter?

- *benefits, challenges, recommendations for improvement*

Wrap-up

- Are there questions that I might have overlooked in asking?
- Do you have any other inputs?
- Thank you for taking part in this in-depth interview!
