## Supplementary material for "Optimizing delivery strategies for 3HP TB preventive treatment in Tanzania: A qualitative study on acceptability of family approach in HIV care and treatment centers": In-depth interview guides: S2_In-depth interview guide_Community health workers.docx

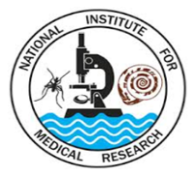
 **IN-DEPTH INTERVIEW GUIDE (CHW): English**

**Interviewer Information**

Name of Interviewer:

Location:

Interviewee code:

Date:

**Introductory Remarks:**

Thank you for consenting to participate in this in-depth interview. I am.......................... and I would like to discuss with you about your views and experiences in the implementation of the family approach on the use of Tuberculosis preventive treatment (TPT) as prophylaxis against TB disease. In particular, I would like to understand the challenges you have experienced and ways you have been able to cope with them.

**Discussion Topics**

1. I would like to start by collecting some demographic information: sex, education level, occupation, facility and years of experience as a CHW

**Knowledge about family approach to TPT provision (Intervention coherence)**

2. What do you understand about Tuberculosis preventive treatment?

- *uses of TPT*
- *duration*
- *who should take it?*
- *dosing, accessibility*
- *ongoing challenges in its uptake among PLHIV*

3. Please tell me what you know about using a family approach to TPT provision

- *rationale*
- *procedures involved*
- *how was information obtained?*
- *stakeholders involved*

**Confidence in performing the behavior required for family approach to TPT**

**(Self-efficacy)**

4. To what extent did you view your ability to implement household HIV testing among household members of the PLHIV?

- *anticipated barriers*
- *confidence in HIV counselling*
- *confidence in successful linkage to care*

**Capacity of family approach to increase uptake of TPT (Perceived effectiveness)**

5. In your opinion, what can hinder the ability of using treatment supporters to support the use of TPT among PLHIV?

- *busy lifestyle*
- *impatience*
- *uninterested*
- *lack of support skills*

6. What are your thoughts about the use of SMS reminders in supporting TPT adherence and in identifying TB symptoms?

- *challenges, recommendations for improvement*

7. What could hinder health providers to provide better services for TPT during clinic consultations?

- *patient workload*
- *unfriendly language*
- *inadequate counselling (privacy)*

**Feelings about receipt of TPT via family approach (Affective attitude)**

8. When you think about the way in which TPT is offered under the support of a treatment supporter and your role in household testing and linkage to care so far, what aspects do you like?

- *health education sessions at the clinic*
- *use of treatment supporters, household HIV testing*

9. When you think about the way in which TPT is offered under the support of a treatment supporter and your role in household testing and linkage to care so far, what are the aspects that you do not like?

- *health education sessions at the clinic*
- *use of treatment supporters, household HIV testing*

**Intervention fitness with lifestyle and work responsibilities (Ethicality)**

10. In your opinion, has household HIV testing in addition to TB screening influenced changes in your work burden?

- *changes in work schedule*

**Efforts/challenges experienced in supporting TPT via the family approach (Burden/opportunity costs)**

11. What barriers did you experience when reaching out for your household members for HIV testing and TB screening?

- *unwillingness of household members*
- *privacy issues*
- *unwillingness of linkage to care*

12. What aspects of your tasks have proven to be challenging?

- *HIV counselling, TB screening, linkage to care*
- *coping strategies*
- *kind of support needed*

Wrap-up

- Are there questions that I might have overlooked in asking?
- Do you have any other inputs?
- Thank you for taking part in this in-depth interview!
