## Supplementary material for "Optimizing delivery strategies for 3HP TB preventive treatment in Tanzania: A qualitative study on acceptability of family approach in HIV care and treatment centers": In-depth interview guides: S3_Topic guide_Treatment supporters.docx

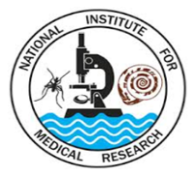
**TOPIC GUIDE (TREATMENT SUPPORTERS)**

**FGD Information**

Name of Facilitator:

Name of Note Taker:

Location:

Focus Group Discussion code:

Date:

**Introductory Remarks:**

Thank you for consenting to participate in this focus group discussion. I am.......................... and my colleague is………………We would like to discuss with the team about your views and experiences in providing support for TPT use among PLHIV.

**Knowledge about family approach to TPT provision (Intervention coherence)**

1. Please tell me what you understand about Tuberculosis

- *transmission*
- *high risk groups*
- *treatment*
- *prevention (prophylaxis)*

2. What do you understand about Tuberculosis preventive treatment?

- *uses of TPT*
- *duration*
- *who should take it?*
- *dosing, accessibility*
- *what do others say about it?*

3. Please tell me what you know about using a family approach to TPT provision

- *rationale*
- *procedures involved*
- *how was information obtained?*
- *stakeholders involved*

**Confidence in performing the behavior required for family approach to TPT**

**(Self-efficacy)**

4. What motivated you to accept providing support for TPT adherence to the PLHIV?

- *TB risk perception, influence from others*

5. How did you foresee your ability to support the PLHIV throughout the 6 months period?

- *anticipated challenges*

**Capacity of family approach to increase uptake of TPT (Perceived effectiveness)**

6. In your opinion, what can hinder the ability of using treatment supporters to support the use of TPT among PLHIV?

- *busy lifestyle*
- *impatience*
- *uninterested*
- *lack of support skills*

7. How has weekly SMS reminders helped you in supporting TPT adherence for the PLHIV and in identifying his/her TB symptoms?

- *more attentive*

8. What could hinder health providers to provide better services for TPT during clinic consultations and health education sessions?

- *patient workload*
- *unfriendly language*
- *inadequate counselling (privacy)*

**Feelings about receipt of TPT via family approach (Affective attitude)**

9. When you think about the way in which TPT is offered to PLHIV, from how you were approached to the support you have been giving so far, what aspects do you like?

- *health education sessions*
- *SMS reminders*
- *support from CHWs*

10. When you think about the way in which TPT is offered to PLHIV, from how you were approached to the support you have been giving so far, what are the aspects that you do not like?

- *health education sessions*
- *SMS reminders*
- *support from CHWs*

**Intervention fitness with lifestyle and work responsibilities (Ethicality)**

11. In your opinion, has the support that you have been providing to the PLHIV changed the way you live or work?

- *changes in work schedule, anxiety*

**Efforts/challenges experienced in supporting TPT via the family approach (Burden/opportunity costs)**

12. What opinions do you have about participating in health education sessions at the clinic with your PLHIV?

- *benefits, challenges, recommendations for improvement*

13. What challenges have you been experiencing in supporting TPT adherence to the PLHIV?

- *coping strategies*

Wrap-up

- Do you have any other inputs?
- Thank you for taking part in this in-depth interview!

**Focus Group Discussion (Treatment supporters)**

**Demographics**

**Participant Information**

Date: ________________________________

FGD number:____________________________

Sex: ­­­­­­­­­­­­­­­­­____________________________

Education level: ­­­­­­­­­­­­­­­­­____________________________

Occupation:________________________________

Name of facility of PLHIV: _____________________________________
